# Antidepressant drug prescription and incidence of COVID-19: a retrospective cohort study

**DOI:** 10.1101/2022.12.15.22283507

**Authors:** Oleg O. Glebov, Christoph Mueller, Robert Stewart, Dag Aarsland, Gayan Perera

## Abstract

While antidepressant drugs (ADs) have shown some efficacy in treatment of COVID-19, their preventative potential remains unexplored. To investigate association between AD and COVID-19 incidence in the community, we analysed data from community-living, non-hospitalized adults admitted to inpatient care of the South London&’Maudsley (SLaM) NHS Foundation Trust during the 1st wave of COVID-19 pandemic in the UK. Prescription of ADs within the period of 1 to 3 months before admission was associated with an approximately 40% decrease in positive COVID-19 test results when adjusted for socioeconomic parameters and physical health. This association was specifically observed for ADs of the Selective Serotonin Reuptake Inhibitor (SSRI) class. These results suggest that ADs, specifically SSRIs, may help prevent COVID-19 infection in the community. Definitive determination of AD preventative potential warrants prospective studies in the wider general population.

**Key points:** *Question:* Is there an association between prescription of antidepressants and incidence of COVID-19 in the general population?

*Findings:* In this retrospective cohort study of mental health outpatients with a recent (1-3 months) antidepressant prescription, there was a statistically significant 40% decrease in positive COVID-19 tests. This association was specifically observed for the most commonly prescribed antidepressant class, Selective Serotonin Reuptake Inhibitors, and remained when adjusted for socioeconomic parameters and physical health.

*Meaning:* Antidepressant prescription was associated with lower incidence of COVID-19 in the community, warranting further investigation as prophylactics in prospective clinical studies.

## Main text

More than 2.5 years since the declaration of the global COVID-19 pandemic, it remains a major public health burden across the world, with further waves of infection more likely than not. To this day, the only available method for prevention of COVID-19 is vaccination. Although development of vaccines has to some extent limited the spread of the SARS-CoV-2 infection, their production, storage and distribution remain a formidable logistical challenge, which is further compounded by the need for repeated administration^1^. There is also persistent uncertainty regarding the safety profile and long-term effects of vaccination, as well as concerning the efficacy of existing vaccines against newly emerging SARS-CoV-2 variants^2,3^. Moreover, vaccines offer limited protection for immunocompromised individuals^4^. Taken together, these considerations highlight a significant unmet need for development of alternative strategies for mitigating the risk and impact of COVID-19.

One potentially promising approach involves repurposing well-characterised existing drugs. Early high-profile failures of hydroxychloroquine^5^ and ivermectin^6^ notwithstanding, more recently it has been shown that antidepressant drugs (AD) may be associated with improved outcomes in hospitalised COVID-19 patients^7^; furthermore, one AD (fluvoxamine) has shown efficacy in preventing severe COVID-19 in clinical trials^8–11^, and another (fluoxetine) was associated with a slight decrease in mortality in a large cohort of COVID-19 patients^12^. To date, however, the effectiveness of ADs for COVID-19 prevention has not been assessed.

Prior cell biological evidence indicates that ADs may affect membrane trafficking^13,14^; on this basis, we previously postulated that subversion of membrane trafficking of the host cell provides a plausible mechanism for COVID-19 protection^15^, and we confirmed that low concentrations of fluvoxamine modulate trafficking of SARS-CoV-2 Spike protein in human cells^16^. To investigate a potential link between ADs and COVID-19 protection *in vivo*, we analysed associations between incidence of positive COVID-19 test results and prior AD prescription, using clinical data derived from individuals admitted to the inpatient units of the South London & Maudsley (SLaM) NHS Foundation Trust^17^ during the 1st phase of the UK’s COVID-19 pandemic.

We leveraged data from the Clinical Record Interactive Search (CRIS) platform, which provides research access to deidentified electronic clinical records for SLaM^18^. During the study period (1 April -31 December 2020), 5664 cases of mental health inpatient care admission at SLaM facilities had been tested for COVID-19, with 202 (3.56%) testing positive. Patients were also assessed on HoNOS (Health of the Nation Outcome Scales), which is a standard measure of health and social functioning used by British mental health services^19,20^. Characteristics of the study cohort are presented in **Table 1**. By ICD-10 code, the most prevalent primary diagnoses were in the F2 (schizophreniform) category, and the second most common in the F3 (mood disorders) category.

**Table 1:**
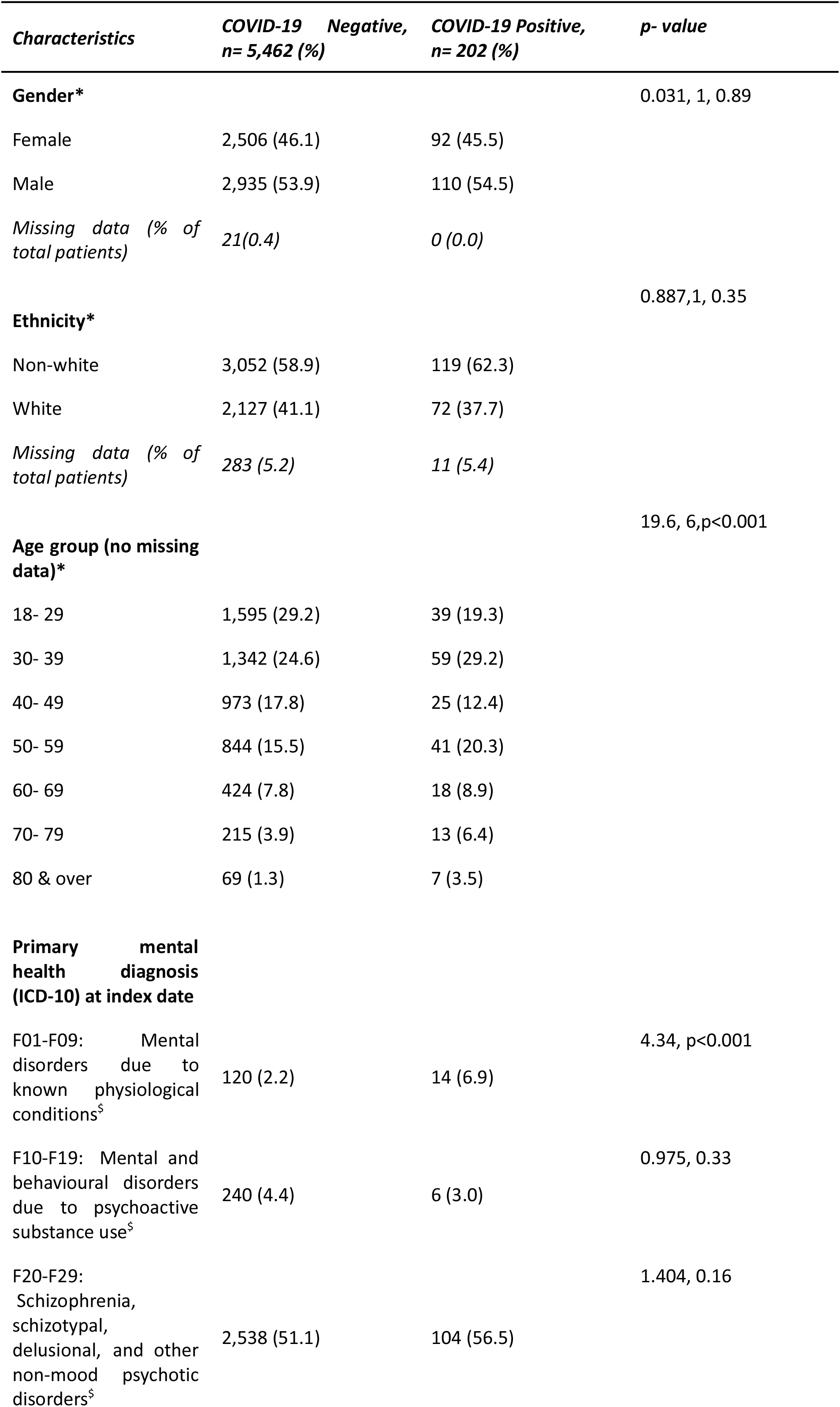
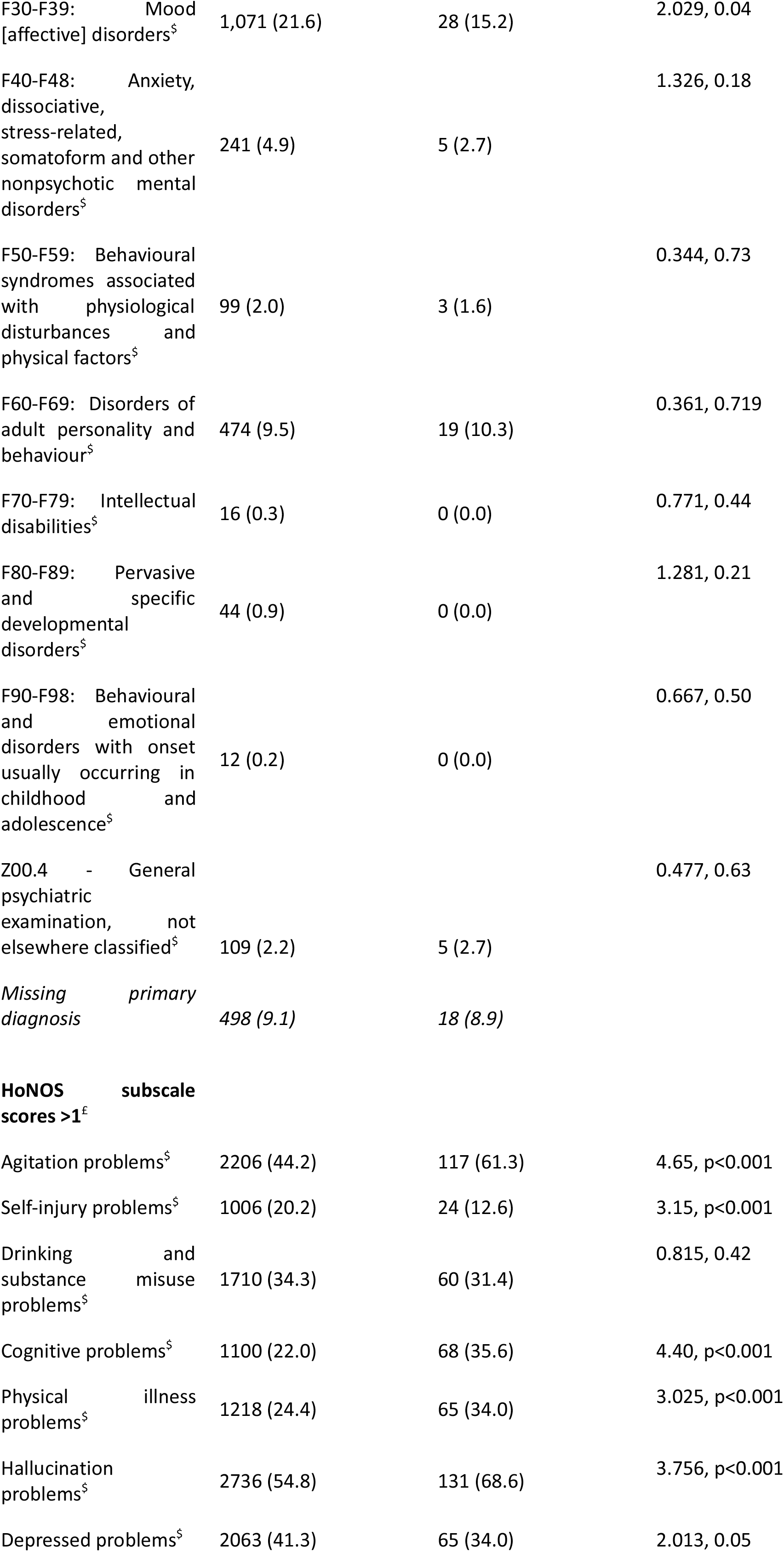
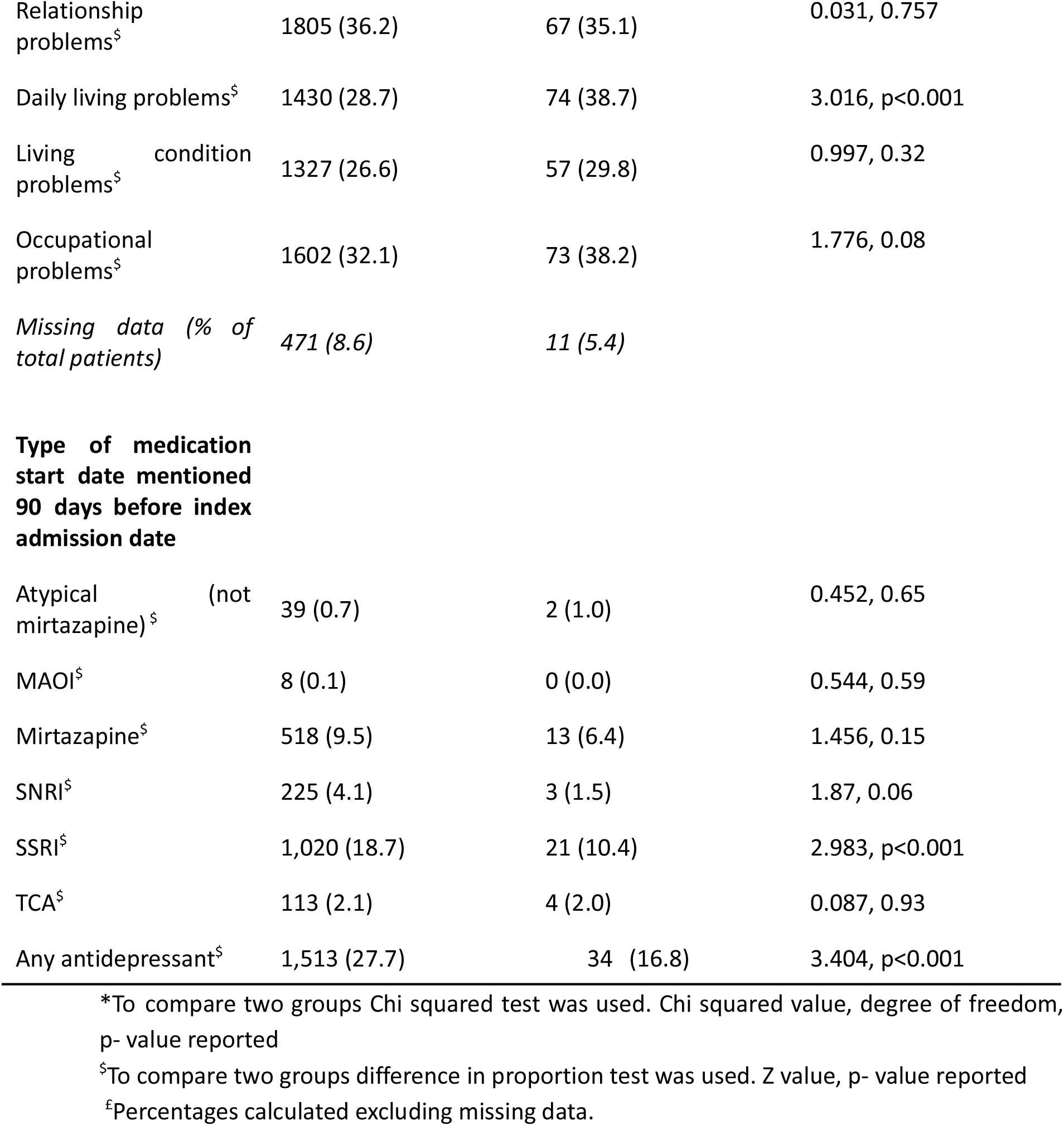
Characteristics of the study cohort

| **Characteristics** | **COVID-19 Negative, n= 5,462 (%)** | **COVID-19 Positive, n= 202 (%)** | **p- value** |
| --- | --- | --- | --- |
| **Gender\*** |  |  | 0.031, 1, 0.89 |
| Female | 2,506 (46.1) | 92 (45.5) |  |
| Male | 2,935 (53.9) | 110 (54.5) |  |
| Missing data (% of total patients) | 21(0.4) | 0 (0.0) |  |
| **Ethnicity\*** |  |  | 0.887,1, 0.35 |
| Non-white | 3,052 (58.9) | 119 (62.3) |  |
| White | 2,127 (41.1) | 72 (37.7) |  |
| Missing data (% of total patients) | 283 (5.2) | 11 (5.4) |  |
| **Age group (no missing data)\*** |  |  | 19.6, 6,p<0.001 |
| 18- 29 | 1,595 (29.2) | 39 (19.3) |  |
| 30- 39 | 1,342 (24.6) | 59 (29.2) |  |
| 40- 49 | 973 (17.8) | 25 (12.4) |  |
| 50- 59 | 844 (15.5) | 41 (20.3) |  |
| 60- 69 | 424 (7.8) | 18 (8.9) |  |
| 70- 79 | 215 (3.9) | 13 (6.4) |  |
| 80 & over | 69 (1.3) | 7 (3.5) |  |
| **Primary mental health diagnosis (ICD-10) at index date** |  |  |  |
| F01-F09: Mental disorders due to known physiological conditions<sup>§</sup> | 120 (2.2) | 14 (6.9) | 4.34, p<0.001 |
| F10-F19: Mental and behavioural disorders due to psychoactive substance use<sup>§</sup> | 240 (4.4) | 6 (3.0) | 0.975, 0.33 |
| F20-F29: Schizophrenia, schizotypal, delusional, and other non-mood psychotic disorders<sup>§</sup> | 2,538 (51.1) | 104 (56.5) | 1.404, 0.16 |
| F30-F39: Mood<br>[affective] disorders<sup>§</sup> | 1,071 (21.6) | 28 (15.2) | 2.029, 0.04 |
| F40-F48: Anxiety,<br>dissociative,<br>stress-related,<br>somatoform and other<br>nonpsychotic mental<br>disorders<sup>§</sup> | 241 (4.9) | 5 (2.7) | 1.326, 0.18 |
| F50-F59: Behavioural<br>syndromes associated<br>with physiological<br>disturbances and<br>physical factors<sup>§</sup> | 99 (2.0) | 3 (1.6) | 0.344, 0.73 |
| F60-F69: Disorders of<br>adult personality and<br>behaviour<sup>§</sup> | 474 (9.5) | 19 (10.3) | 0.361, 0.719 |
| F70-F79: Intellectual<br>disabilities<sup>§</sup> | 16 (0.3) | 0 (0.0) | 0.771, 0.44 |
| F80-F89: Pervasive<br>and specific<br>developmental<br>disorders<sup>§</sup> | 44 (0.9) | 0 (0.0) | 1.281, 0.21 |
| F90-F98: Behavioural<br>and emotional<br>disorders with onset<br>usually occurring in<br>childhood and<br>adolescence<sup>§</sup> | 12 (0.2) | 0 (0.0) | 0.667, 0.50 |
| Z00.4 - General<br>psychiatric<br>examination, not<br>elsewhere classified<sup>§</sup> | 109 (2.2) | 5 (2.7) | 0.477, 0.63 |
| *Missing primary*<br>*diagnosis* | *498 (9.1)* | *18 (8.9)* |  |
| **HoNOS subscale**<br>**scores >1<sup>£</sup>** |  |  |  |
| Agitation problems<sup>§</sup> | 2206 (44.2) | 117 (61.3) | 4.65, p<0.001 |
| Self-injury problems<sup>§</sup> | 1006 (20.2) | 24 (12.6) | 3.15, p<0.001 |
| Drinking and<br>substance misuse<br>problems<sup>§</sup> | 1710 (34.3) | 60 (31.4) | 0.815, 0.42 |
| Cognitive problems<sup>§</sup> | 1100 (22.0) | 68 (35.6) | 4.40, p<0.001 |
| Physical illness<br>problems<sup>§</sup> | 1218 (24.4) | 65 (34.0) | 3.025, p<0.001 |
| Hallucination<br>problems<sup>§</sup> | 2736 (54.8) | 131 (68.6) | 3.756, p<0.001 |
| Depressed problems<sup>§</sup> | 2063 (41.3) | 65 (34.0) | 2.013, 0.05 |
| Relationship problems<sup>§</sup> | 1805 (36.2) | 67 (35.1) | 0.031, 0.757 |
| Daily living problems<sup>§</sup> | 1430 (28.7) | 74 (38.7) | 3.016, p<0.001 |
| Living condition problems<sup>§</sup> | 1327 (26.6) | 57 (29.8) | 0.997, 0.32 |
| Occupational problems<sup>§</sup> | 1602 (32.1) | 73 (38.2) | 1.776, 0.08 |
| *Missing data (% of total patients)* | *471 (8.6)* | *11 (5.4)* |  |
| **Type of medication start date mentioned 90 days before index admission date** |  |  |  |
| Atypical (not mirtazapine)<sup>§</sup> | 39 (0.7) | 2 (1.0) | 0.452, 0.65 |
| MAOI<sup>§</sup> | 8 (0.1) | 0 (0.0) | 0.544, 0.59 |
| Mirtazapine<sup>§</sup> | 518 (9.5) | 13 (6.4) | 1.456, 0.15 |
| SNRI<sup>§</sup> | 225 (4.1) | 3 (1.5) | 1.87, 0.06 |
| SSRI<sup>§</sup> | 1,020 (18.7) | 21 (10.4) | 2.983, p<0.001 |
| TCA<sup>§</sup> | 113 (2.1) | 4 (2.0) | 0.087, 0.93 |
| Any antidepressant<sup>§</sup> | 1,513 (27.7) | 34 (16.8) | 3.404, p<0.001 |

\*To compare two groups Chi squared test was used. Chi squared value, degree of freedom, p- value reported
<sup>§</sup>To compare two groups difference in proportion test was used. Z value, p- value reported
<sup>£</sup>Percentages calculated excluding missing data.

We then queried CRIS for mentions of ADs in the clinical records of patients within the time period of 90 days preceding the date of admission. The list of drugs included in the query is presented in **Table 2**. 27.7% percent of COVID-19-negative cases had at least one AD mention within 90 days preceding admission, compared to 16.8% of COVID-19-positive cases. Accordingly, the occurrence of a positive COVID-19 test result in patients with an AD mention was significantly lower than in those without (2.2 *vs* 4.1%, p = .000663, X^2^ test). Most prescribed ADs belonged to the Selective Serotonin Reuptake Inhibitor (SSRI) class: two thirds (67%) of the cases with AD mention within 90 days (**Table 1**), and the occurrence of a COVID-19-positive test result in patients with a recent SSRI record was significantly less than in those without (2.0 *vs* 3.9%, p = .002853, X^2^ test). Associations with other AD classes were not statistically significant.

**Table 2:** Drugs assessed in this study

| ***Drug class*** | ***Drug name*** |
| --- | --- |
| Selective Serotonin Reuptake Inhibitor (SSRI) | citalopram<br>dapoxetine<br>escitalopram<br>fluoxetine<br>fluvoxamine<br>paroxetine<br>sertraline |
| Serotonin and Norepinephrine Reuptake Inhibitor (SNRI) | venlafaxine<br>duloxetine<br>reboxetine |
| Atypical | mirtazapine<br>vortioxetine<br>bupropion<br>trazodone<br>vilazodone |
| Tricyclic antidepressant (TCA) | amitriptyline<br>clomipramine<br>imipramine<br>lofepramine<br>nortriptyline<br>trimipramine |
| Monoamine Oxidase Inhibitor (MAOI) | tranylcypromine<br>phenelzine<br>isocarboxazid<br>moclobemide |

To further investigate the relationship between AD/SSRI and COVID-19 test results, we performed multiple logistic regression analysis (**Table 3)**. We found significant adjusted associations between AD receipt within 31, 60, and 90 days before admission and COVID-19 test results. Similar associations were found for SSRI receipt within 62 and 90 days prior to admission in adjusted models. Following stratification by diagnosis (**Table 4**), associations were similar in direction across all groups apart from substance use disorders, and were strongest in organic disorders (F0x), mood disorders (F3x) and anxiety disorders (F4x).

**Table 3:** Strength of the association between any antidepressants medication receiving status and COVID-19 positive test result at when medication was mentioned 31 days, 62 days and 90 days for the first time before index hospital admission [Odds ratios (95% CI), P value]. Statistically significant association in bold.

| *Type of medication* | *Adjustments* | *31 days* | *62 days* | *90 days* |
| --- | --- | --- | --- | --- |
| Any drug received | unadjusted | **0.52 (0.34, 0.81), <0.001** | **0.51 (0.35, 0.76), <0.001** | **0.53 (0.36, 0.77), <0.001** |
|  | adjusted for sociodemographic factors (gender, age, ethnicity) | **0.55 (0.35, 0.86), 0.01** | **0.52 (0.35, 0.79), <0.001** | **0.54 (0.37, 0.80), <0.001** |
|  | *Further adjusted for HoNoS symptoms and primary mental health diagnosis) | **0.59 (0.36, 0.96), 0.03** | **0.61 (0.39, 0.95), 0.03** | **0.61 (0.39, 0.94), 0.02** |
| SSRIs | unadjusted | **0.52 (0.30, 0.90), 0.02** | **0.50 (0.31, 0.8), <0.001** | **0.51 (0.32, 0.80), <0.001** |
|  | adjusted for sociodemographic factors (gender, age, ethnicity) | 0.59 (0.34, 1.03), 0.06 | **0.53 (0.32, 0.87), 0.01** | **0.55 (0.34, 0.88), 0.01** |
|  | *Further adjusted for HoNoS symptoms and primary mental health diagnosis) | 0.59 (0.31, 1.12), 0.10 | **0.54 (0.30, 0.95), 0.03** | **0.57 (0.32, 0.95), 0.03** |
| SNRIs | unadjusted | 0.16 (0.02, 1.14), 0.07 | 0.25 (0.06, 1.02), 0.05 | 0.35 (0.11, 1.11), 0.07 |
|  | adjusted for sociodemographic factors (gender, age, ethnicity) | 0.16 (0.02, 1.12), 0.07 | 0.25 (0.06, 1.02), 0.05 | 0.35 (0.11, 1.11), 0.08 |
|  | *Further adjusted for HoNoS symptoms and primary mental health diagnosis) | 0.23 (0.03, 1.66), 0.15 | 0.38 (0.09, 1.37), 0.10 | 0.53 (0.16, 1.63), 0.24 |
| Mirtazapine | unadjusted | 0.73 (0.38, 1.39), 0.34 | 0.65 (0.36, 1.18), 0.16 | 0.66 (0.37, 1.16), 0.15 |
|  | adjusted for sociodemographic factors (gender, age, ethnicity) | 0.71 (0.37, 1.38), 0.32 | 0.64 (0.35, 1.16), 0.14 | 0.65 (0.36, 1.15), 0.14 |
|  | *Further adjusted for HoNoS symptoms and primary mental health diagnosis) | 1.01 (0.51, 2.01), 0.97 | 0.94 (0.50, 1.57), 0.64 | 0.97 (0.52, 1.77), 0.91 |

\*Adjusted for all significant HoNoS subscale and primary mental health diagnosis variables in the table 1 (F01-F09: Mental disorders due to known physiological conditions, F30-F39: Mood [affective] disorders, Agitation problems, Self-injury problems, Cognitive problems, Physical illness problems, Hallucination problems, Depressed problems, Daily living problems

**Table 4:**
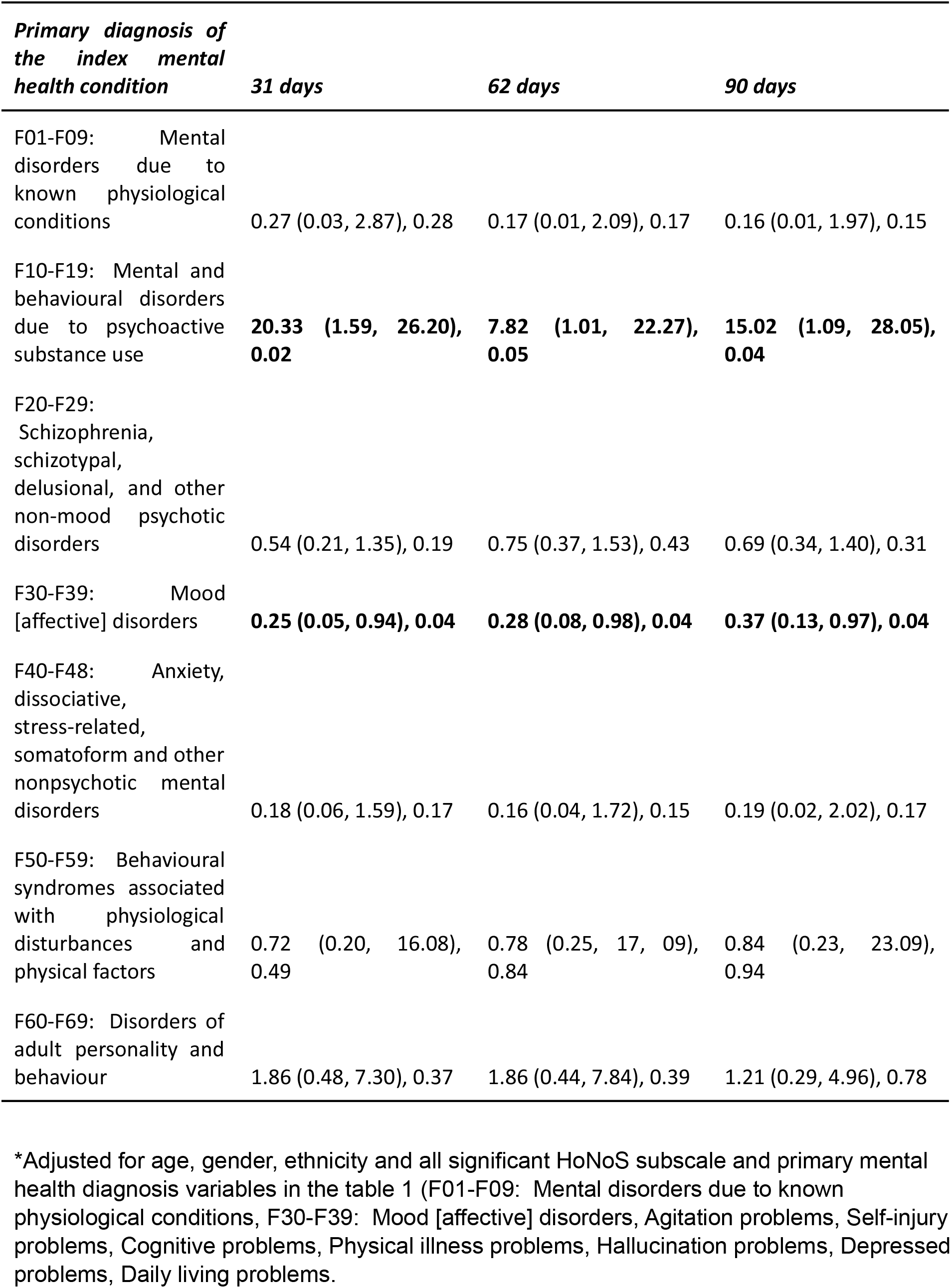
Strength of the association between any antidepressants medication receiving status and COVID-19 positive test result when medication was mentioned at 90 days for the first time before index hospital admission [Odds ratios (95% CI), P value] stratified by primary mental health condition at the time of index hospital admission. Statistically significant association in bold.

The results of this study indicate that prior receipt of ADs and specifically SSRIs was associated with a decreased likelihood of COVID-19 incidence in people receiving mental health inpatient care, suggesting that these drugs may have a protective effect against COVID-19 in this population. One key strength of this study is the large number of participants. Furthermore, the focus on the 1st wave of COVID-19 avoids the confounding effects of the “herd immunity” in the population due to mass vaccination and/or previous exposure to COVID-19 in the following time period^21^. Importantly, exposure to both ADs and COVID-19 occurred prior to hospitalisation, reflecting the “real-life” situational value of the study, which is further enhanced by the diverse socio-economical composition of the study cohort (**Table 1)**.

The study has a number of important limitations, largely due to its retrospective nature and focus on a specific cohort of mental health inpatients. It cannot demonstrate a causal relationship between medications and COVID-19 test results; indeed, the protective effect may be due to either decreased infection rate or increased recovery rate from COVID-19. Owing to the numbers of participants and multiple drug prescriptions, it was not possible to investigate the association between COVID-19 and individual drugs; compared to SSRIs, the number of participants receiving other drug classes was low (**Table 1**), and it was also not possible to assess older-generation drugs, including fluvoxamine. Finally, although the association of interest appeared to be present across a range of diagnostic groups, it is possible that antidepressant use was a marker of other personal or clinical factors conferring protection. To overcome the above limitations and to corroborate the findings of this study, randomised prospective clinical trials for selected ADs in the general population will be necessary.

SSRI treatment is associated with a dropout rate of 28% over the standard 6-month treatment course^22^. Given the study design, it can be expected that a proportion of cases did not adhere to the prescription, suggesting that the observed effect may be an under-estimation, Nevertheless, the reduction in hazard ratio reported here is considerably larger that 8% reduction in overall mortality associated with record of SSRI in a large cohort of COVID-19 cases^12^, being more similar to the 36% reduction by fluvoxamine in risk of hospitalisation for outpatients with COVID-19^11^, and to the hazard ratio decrease of 44% reported for intubated hospitalised patients on SSRIs^7^. In the context of the above studies, our findings indicate that AD/SSRIs may be at least as effective in preventing COVID-19 as in treating it, providing impetus for further investigation of their clinical utility in the general population.

To conclude, our results suggest that ADs, and SSRIs in particular, may protect against COVID-19. This evidence lends support at least to further investigation of drug repurposing as a viable alternative to vaccination, especially considering the key advantages of ADs over the recently developed vaccines, *viz*. well-characterised safety profile, low price and high stability. Identification of affordable and safe drugs that reduce the risk of or from COVID-19 infection is likely to be important for global pandemic responses, especially in cases and situations where vaccination -especially mass vaccination -may be problematic. In the longer term, it may be worth investigating the utility of ADs/SSRIs for treatment of respiratory infections that may use similar cell biological mechanisms to COVID-19, *e*.*g*. influenza^23^. For now, we hope that the results from this study will help inform the public health policy debate on COVID-19 management, re-establish drug repurposing in the context of COVID-19 treatment and highlight the potential for wider clinical benefits of psychotropic drugs.

## Methods

### Study design, data source, and population

We conducted an observational, retrospective, matched cohort study of individuals admitted to the 4 inpatient care units of South London and Maudsley (SLaM) NHS Foundation Trust in the 1st wave of the COVID-19 pandemic of 2020. The inpatient care units affiliated with SLAM are as follows: Bethlem Royal Hospital, Lambeth Hospital, Lewisham Hospital, and Maudsley Hospital^17^. SLaM provides near-monopoly comprehensive mental health services to a geographic catchment of 1.3m residents in four boroughs of south London. SLaM has used electronic health records across all its services since 2006 and its Clinical Record Interactive Search (CRIS) platform was set up in 2008 to provide researcher access to deidentified data from these records within a robust governance infrastructure. CRIS has been subsequently developed through a range of data linkages and natural language processing (NLP) algorithms^18^, and the platform has supported over 250 peer-reviewed publications to date. Using CRIS we extracted data on cases admitted to a SLaM inpatient facility between 1 April and 31 December 2020, and who were aged 18 years or over at the time of admission. A PCR or antigen test for COVID-19 was routinely performed at admission and during the inpatient stay over that period. The criterion for inclusion was the conclusive positive or negative COVID-19 test result(s) during inpatient stay in a SLaM inpatient unit. Characteristics of the study cohort are presented in **Table 1**.

### Exposure & Outcome

We used an NLP algorithm to identify the medications mentioned in the patient’s record in a 6-month window before or after referral 28. The list of medications can be found in Table 2. We established use of the following medication classes: atypical (not mirtazapine), monoamine oxidase inhibitors (MAOI), mirtazapine, serotonin & norepinephrine reuptake inhibitors (SNRI), selective serotonin reuptake inhibitors (SSRI), tricyclic antidepressants (TCA) and any antidepressants mentioned above within 31, 62, or 90 days preceding the index hospital admission.

The primary outcome variable was the result of the laboratory test for COVID-19 (antigen or PCR). Any incidence of a positive COVID-19 test result during the inpatient stay was categorised as “positive”. For categorization as “negative” all COVID test results during inpatient stay were required to be negative.

### Covariates & Predictors

We ascertained age at the time of admission, gender, and ethnicity (dichotomised to White and non-White) as recorded at the time of hospital admission. We identified the diagnosis given closest to hospital admission in structured fields. According to routinely applied WHO ICD-10^24(p10)^ criteria for diagnostic codings in the record, we established the following diagnosis groups:

F01-F09: Mental disorders due to known physiological conditions;

F10-F19: Mental and behavioural disorders due to psychoactive substance use;

F20-F29: Schizophrenia, schizotypal, delusional, and other non-mood psychotic disorders;

F30-F39: Mood [affective] disorders;

F40-F48: Anxiety, dissociative, stress-related, somatoform and other nonpsychotic mental disorders;

F50-F59: Behavioural syndromes associated with physiological disturbances and physical factors;

F60-F69: Disorders of adult personality and behaviour;

F70-F79: Intellectual disabilities;

F80-F89: Pervasive and specific developmental disorders;

F90-F98: Behavioural and emotional disorders with onset usually occurring in childhood and adolescence.

We established mental and physical health problems, as well as functional difficulties using the Health of the Nation Outcome Scales (HoNOS)^20^. This is a routinely used measure in British mental health services and the most recent scores were extracted at the time of the index admission. Each subscale is rated on a scale ranging from 0 (no problem) to 4 (severe or very severe problem); to simplify interpretation, we dichotomised the scores to “minor or no problems” (scores 0 or 1) and ‘mild to severe problems’ (scores 2 to 4).

### Statistical techniques

Initially, chi squared tests and difference in proportion statistical test using Z-values for difference in two groups between patients receiving COVID-19 positive test and COVID-19 negative results were tested for each covariate. Logistic regression models were assembled to quantify odds ratios (ORs) for the associations between antidepressant medication receipt and COVID-19 positive test result, applying the above sub-categorisation of antidepressants and timing as secondary analyses. 95% confidence intervals (CI) and P-values for ORs were calculated, and P-values <0.05 were considered statistically significant. Initially unadjusted logistic regression analyses were carried out, followed by adjustments for sociodemographic factors (gender, age, ethnicity) and then further adjustments for those significant HoNoS subscales and primary mental health diagnosis. Finally, primary analyses (only patients receiving any antidepressant medication) were stratified by primary mental health diagnosis measured using ICD-10 diagnosis at the time of index hospital admission. All statistical analyses were conducted with STATA version15^25^.

## Data Availability

All data produced in the present work are contained in the manuscript

## Authors’ Contributions

Study concept: OOG, CM, DA

Study design: OOG, CM, GP

Project administration: OOG, CM, RS, DA, GP

Data extraction and verification: CM, RS, GP

Data analysis: OOG, GP

Manuscript writing and revisions: OOG, CM, RS, DA, GP

## Funding

OOG is funded by the Lewy Body Society and the National Natural Science Fund of China. RS, CM, DA and GP are part-funded by the National Institute for Health Research (NIHR) Biomedical Research Centre at the South London and Maudsley NHS Foundation Trust and King’s College London. RS is additionally part-funded by the National Institute for Health Research (NIHR) Applied Research Collaboration South London (NIHR ARC South London) at King’s College Hospital NHS Foundation Trust, and the DATAMIND HDR UK Mental Health Data Hub (MRC grant MR/W014386). The views expressed are those of the authors and not necessarily those of the NIHR or the Department of Health and Social Care.

**Figure 1.**
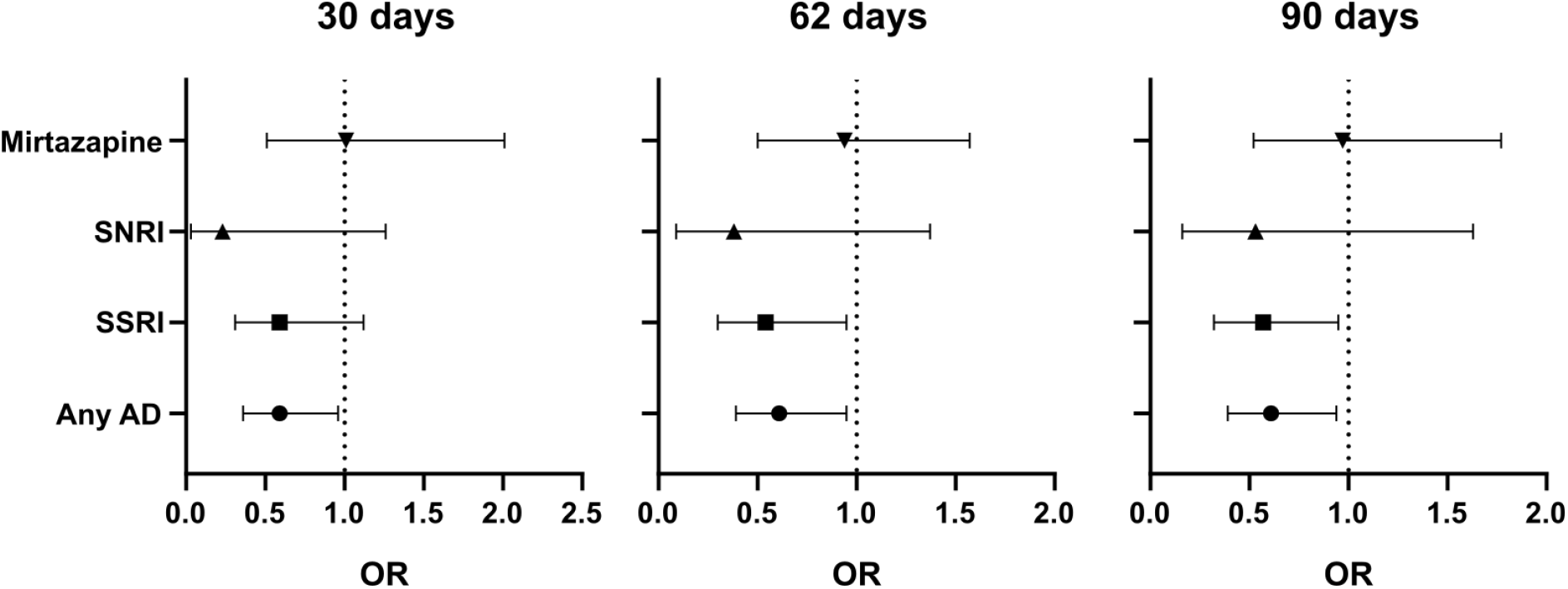
Adjusted Odds Ratios (OR) at 30, 62, and 90 days (related to Table 3). Error bars correspond to 95% CI.

